# Feasibility, Safety and Efficacy of OsteoStrong^®^ in Postmenopausal Women with Low Bone Mineral Density: A Pilot Study

**DOI:** 10.1101/2025.05.23.25328203

**Authors:** Jakub Mesinovic, Jack R Ryan, Carrie-Anne Ng, Ayse Zengin, Peter R Ebeling, David Scott

## Abstract

OsteoStrong^®^ is described as a high-intensity, low-volume exercise modality that targets bone health improvements by applying isometric forces at relevant anatomical sites using specific equipment during brief (∼10-minute, once weekly) sessions. Evidence on adherence, safety and effectiveness is limited. In this single-arm interventional study, we determined the feasibility of 8 months of OsteoStrong^®^ for postmenopausal women with low BMD, and measured changes in bone density, microarchitecture and strength, physical function and body composition. Forty-four postmenopausal women with low BMD (DXA-determined T-score <-1.0 but >-3.0 at total hip and/or lumbar spine) attended supervised, once-weekly 10–15-minute sessions at an OsteoStrong^®^ clinic for 8 months. We calculated 8-month changes in areal BMD (aBMD), trabecular bone scores (TBS), HRpQCT-determined volumetric BMD (vBMD) and bone microarchitecture and strength, as well as physical function and body composition. Thirty-eight women aged 61.2±5.5 years completed the study. Two adverse events suspected to be intervention-related were recorded. Adherence was 83±28% overall and 93±9% in those who completed the study. At 8 months, there were no changes in total hip, femoral neck, and lumbar spine aBMD (all P>0.05), while TBS decreased (P<0.05). At the distal radius, total, trabecular and cortical vBMD and cortical thickness decreased (mean change: -0.007mm [95%CI: -0.012, -0.002]). At the distal tibia, cortical vBMD decreased and trabecular separation increased (mean change: 0.007mm [95%CI: 0.001, 0.012]). Chair stand time (mean change: -0.8 sec [95%CI: -1.2, -0.5]) and stair climb time (mean change: -0.1 sec [95%CI: -0.2, -0.002]) decreased, while SPPB scores increased (mean change: 0.2 [95%CI: 0.03, 0.38]). There were no changes in other bone, physical function or body composition outcomes. These findings suggest that 8 months of OsteoStrong^®^ does not significantly improve bone density, microarchitecture, or strength in postmenopausal women with low BMD, despite good adherence and safety. However, it improves some measures of physical function.

## 1.0 Introduction

OsteoStrong^®^ is described as a membership-based Integrative Health and Wellness Centre that can generate substantial increases in bone mineral density (BMD), providing benefits for people with poor bone health ^(1)^. OsteoStrong^®^ uses four machines that are claimed to emulate high-impact activity and trigger osteogenesis by applying high-intensity, low-volume, isometric forces to multiple muscle groups and joints in controlled, limited-range movements ^(1)^. The protocol consists of performing a single exercise (i.e., one repetition) on each machine of the patented ‘Spectrum^®^ system’, completed within 10-15 minutes, once per week ^(1)^. As of December 2023, OsteoStrong^®^ comprised almost 200 centres worldwide and reported over 10 million completed client sessions, including 350,000 completed in Australia alone, in the past five years ^(2)^.

At present, only three peer-reviewed studies have published findings on the efficacy of OsteoStrong^®^, so the body of evidence on this program remains limited. A previous study in 55 postmenopausal women reported that 24 weeks of isometric axial bone loading (i.e., delivered via a first-generation OsteoStrong^®^ machine called bioDensity™; four exercises performed once weekly for 15 minutes) increased total hip and lumbar spine areal BMD (aBMD) by ∼15% relative to baseline ^(3)^. However, aBMD was only measured in a subset of the cohort (n=9) and no information on imaging precision was provided ^(3)^. A larger, semi-randomised controlled trial (RCT; The LIFTMOR-M trial) in 93 middle-aged and older men reported that 8 months of the same exercise intervention (bioDensity™; four exercises performed once weekly for 15 minutes) had no effect on total hip, lumbar spine or femoral neck aBMD relative to control (usual activities not involving high-intensity resistance and/or impact training) ^(4)^. Recently, a 12-month controlled study in 140 postmenopausal women utilised the new-generation OsteoStrong^®^ equipment (Spectrum^®^ system), reporting more modest within-group changes in total hip, lumbar spine and femoral neck aBMD (<1.2%) ^(5)^. This is the only study that has evaluated the efficacy of the Spectrum^®^ system used in a commercial setting ^(5)^; however, it provided limited detail on the feasibility of the intervention (particularly with achieving OsteoStrong^®^-determined ‘osteogenic thresholds’) and safety. It also did not report on the precision of imaging outcomes, nor did it utilise advanced three-dimensional imaging techniques capable of detecting volumetric and structural changes in bone that may occur, which are predictive of fractures independently of aBMD ^(6–8)^.

This pilot study aimed to assess adherence to, and safety of OsteoStrong^®^ in postmenopausal women with low aBMD, and to measure changes in bone density and microarchitecture, blood biochemistry, physical function and body composition over 8 months.

## 2.0 Material and Methods

### 2.1 Study design and participants

This 8-month single-arm pilot study recruited 44 postmenopausal women (aged ≥50 years) with low aBMD defined as a dual-energy x-ray absorptiometry (DXA)-determined T-score of <-1.0 at the lumbar spine, or non-dominant total hip or femoral neck. Participants underwent physical screening and baseline assessments at the Monash Health Translational Precinct (MHTP) and returned for follow-up assessments after 8 months. Participants were recruited through print and online advertisements and word of mouth. Following initial phone screening by a research assistant (JRR), potential eligible participants attended the MHTP for further eligibility screening and enrolment into the study. Individuals were eligible if they were community-dwelling, had a body mass index (BMI) <30kg/m² and self-reported <150 minutes of moderate-to-vigorous physical activity (MVPA) per week. Exclusion criteria included a >5% body weight change in the past 3 months, moderate to severe cognitive impairment (Mini-Mental State Exam score <18), osteoarthritis causing severe exercise pain, recent surgeries or injuries affecting the lower or upper limbs, or hernia operations in the past 6 months. Individuals were also excluded if they had dual hip replacement, used medications known to affect bone and mineral metabolism (e.g. bisphosphonates, teriparatide, denosumab etc), or had a DXA T-score ≤-3.0. Individuals with a body mass exceeding 200kg, grip strength <16kg in the preferred hand, reported inability to walk 400 metres unassisted, or a diagnosis of type 1 or type 2 diabetes were also ineligible. A research ethics amendment was approved in July 2022, before the first participant was enrolled, to address recruitment challenges observed where no interested individual was found to be eligible to participate after screening 94 women. This amendment increased the permissible duration for participants to be away from the study centre during the intervention from one to two months, removed requirements for minimum protein and calcium intake, and 25-hydroxyvitamin D (25[OH]D) levels, and allowed women with a myocardial infarction more than 12 months prior to participate with GP approval. These revisions were intended to improve feasibility of recruitment. Participant recruitment and outcome assessment were performed by researchers not affiliated with OsteoStrong^®^. The study was conducted according to the principles of the Declaration of Helsinki, approved by the Monash Health Human Research Ethics Committee (Protocol ID: RES-20-0000-082A), and was prospectively registered with the Australian New Zealand Clinical Trials Registry under ACTRN12620000224921p. All participants provided written informed consent.

### 2.2 Intervention

Participants were provided with a complimentary OsteoStrong^®^ membership for 8 months, entailing supervised, one-on-one training at a single site (OsteoStrong^®^ Hawthorn, Victoria). The regimen prescribed by OsteoStrong^®^ was individually tailored but followed standardised procedures. During this period, exercise intensity and force increased progressively, while the frequency (one day per week) and duration of sessions (approx. 10- 15 minutes) remained constant.

Each OsteoStrong^®^ session began with a whole-body vibration (WBV) warm-up, where participants stood on a Vibeplate (Vibe Plate, Lincoln, NE, USA) vibrating at 30Hz for two minutes. Following the WBV warm-up, participants engaged in four different activities using four machines that comprise the Spectrum^®^ System: the Upper, Lower, Core, and Postural Growth Triggers. Under the supervision of trained OsteoStrong^®^ technicians, participants exerted their perceived maximum force or loading level on each machine without reaching discomfort. These exercises involved limited-range static load contractions with minimal joint angle change, approximately five centimetres in range. Each exertion on a machine lasted 8 to 10 seconds:

1. Upper Growth Trigger (Chest Press): Participants sat on the seat ensuring their back touched the backrest. The technician positioned the bar at the centre of their pectoral muscles, aligned horizontally with their shoulder joint. Gripping the handles with elbows bent at 120 degrees, participants performed a compressive movement similar to a bench press.
2. Lower Growth Trigger (Leg Press): Participants sat on the seat, gripping the handles, and placed their feet shoulder-width apart on the plates with knees bent at 120 degrees. They exerted force through their heels against the plates, resembling a leg press or squat.
3. Core Growth Trigger (Core Pull): Participants sat upright on the seat, used an underhand grip on the handles, and engaged abdominal core muscles in a tucking motion, pulling their arms down and lifting their knees. This exercise combined elements of an underhand pull-up, abdominal crunch, and bent-knee hip flexor exercise.
4. Postural Growth Trigger (Vertical Lift): Participants stood upright between the bars. The technician adjusted the bars to align with their fingertips. Gripping the handles and bending their knees, participants pushed their shoulders back and executed a vertical lift movement, similar to the end range of a deadlift.

Trainers ensured participants maintained a neutral head position at each machine. After a light test movement, they exhaled and exerted maximal force for 8–10 seconds, until the force displayed on the machine screen dropped or muscle fasciculation occurred. Each machine recorded and stored maximum effort data in a central database. An entire circuit lasted 4–6 minutes, with total exertion per participant under one minute. Deconditioned or previously fractured individuals started at 50% of their maximum effort, gradually increasing across sessions to minimise injury risk and enhance conditioning. The remaining participants were encouraged to exert 70–80% of their one-repetition maximum (1RM) in their first session. Each session tracked prior maximum force, motivating participants to exceed or maintain at least 75% of their previous week’s output. Following the exercise circuit, participants underwent another two-minute WBV session on the Vibeplate at 30Hz. OsteoStrong^®^ technicians recorded attendance at each session.

### 2.3 Outcome measures

All outcome measures were collected at baseline and follow-up. Participants completed online questionnaires on demographics, general health and health behaviours. Dietary protein and calcium intake were assessed using 24-hour recalls via the Automated Self-administered Assessment (ASA)-24-Australia tool ^(9)^. Participants recorded their intake over three days (two non-consecutive weekdays and one weekend day) at both time points, listing all foods and beverages consumed, including quantities. Nutrient outliers were identified using cut-off points at the 5th and 95th percentiles, as per ASA24 guidelines.

Participants were asked to wear an ActiGraph wGT3X-BT accelerometer (Actigraph, FL, USA) on their non-dominant wrist (except when completing any water-based activities, i.e. showering, swimming) for seven days. Participants were fitted with the device and given instructions on its use, advised to follow their current lifestyle to obtain representative measurements. Data was analysed using the manufacturer’s software (Version 6.11.2).

Wear and non-wear time was determined using Choi et al ^(10)^ algorithm. Participants were also required to keep a diary to record wear times and reasons for not wearing their device. A trained research assistant (JRR) further verified wear time vs non-wear time data by comparing the algorithm wear-time classifications against diary reports. The accelerometer recorded physical activity data such as steps per day, sedentary time, and moderate and vigorous activity in epoch lengths of 60 seconds. Cut-off points were <2,860 counts/min (sedentary); 2,860-3,940 counts/min (light); and ≥3,941counts/min (moderate-to-vigorous (MVPA) ^(11)^. A valid day was defined as ≥10 hours of wear time during a 24-hour period (00:00 – 23:59 of the same date) ^(12)^. Participants with <4 days consisting of ≥10 hours of wear time were excluded from analyses ^(13)^. Activity records were averaged over valid days to provide a more representative understanding of activity levels.

Except for questionnaires (self-administered) and accelerometer-based measures (devices worn by participants and data analysed by a research assistant [JRR]), almost all data (N=43) were acquired by a single trained individual (JM). Weight was measured to the nearest 0.1kg using an electronic scale (Seca 804, Seca, Germany). Height was measured to the nearest 1mm using a wall-mounted stadiometer (Seca 213, Seca, Germany). Participants were instructed to empty their pockets and remove footwear and heavy clothing prior to measurement.

Participants underwent whole-body DXA scans (Hologic Discovery A, Hologic, USA) for assessment of body composition (appendicular lean mass [ALM; kg] and visceral adipose tissue [VAT; cm^2^]). Scans were also acquired at the total hip and anteroposterior lumbar spine to assess aBMD (g/cm^2^). A lumbar spine trabecular bone score (TBS) was obtained using the TBS iNsight bone texture software (v3.0.2, Medimaps, Switzerland). We used a 4- point scan grading scale to categorise artefacts ^(14)^. A score of 1 was considered perfect. A score of 2 was assigned when a static artefact was present at all time points (e.g., pacemaker, joint replacement). A score of 3 indicated one or two small artefacts present at a single time point (e.g., ring, bra clips). A score of 4 was given when large or multiple small artefacts were present at a single time, rendering the scan poor quality and unusable (e.g., bracelets and zippers). All scans were graded <4 and were acquired and analyzed using the manufacturer’s standard protocol and software (Hologic Apex version 5.6.0.2). CVs for all outcomes have been published elsewhere ^(14)^. The manufacturer’s phantom was scanned on all data collection days for quality control.

Peripheral quantitative computed tomography (pQCT; Stratec XCT3000, Stratec, Germany) scans in the non-dominant tibia. Participants were seated with their tibia positioned inside the pQCT gantry. A single 2.5mm transverse scan was acquired with an in-plane voxel size of 0.5mm and a scan speed of 20mm/sec. Tibia length was measured from the end of the medial malleolus to the tibial plateau. Scan sites were determined using a planar scout view of the distal tibia and a reference line was placed parallel to the distal joint surface of the tibia. Scans were acquired at 66% of the total length of the tibia relative to the reference line. All images were using the manufacturer’s software (version 6.2). CORTBD with separation mode 1 and a threshold of 710 mg/cm³ was used to define cortical volumetric BMD (vBMD) and polar strength-strain index (SSI polar; mm^3^), an estimate of torsional bone strength, was obtained at a threshold of 280 mg/cm³ using cortmode 1. Muscle density (mg/cm^3^) was obtained using a smoothing filter (F03F05) at a threshold of 100 mg/cm^3^. We used a 4-point scan grading scale to categorise motion artefacts in scans ^(14)^. A score of 1 was considered perfect; 2 was considered good with very minor amounts of motion artifact; 3 was assigned to scans where bone data was salvageable but muscle data was not; 4 was given to unusable scans. All scans were graded <4. CVs for all outcomes have been published elsewhere ^(14)^. The manufacturer’s phantom was scanned on all data collection days for quality control.

High-resolution peripheral quantitative computed tomography (HR-pQCT; XtremeCT II, Scanco Medical, Switzerland) scans were acquired in the non-dominant radius and tibia. Limbs were placed in a cast and stabilised to prevent movement artifacts. Scan sites were determined using a planar scout view of the distal radius or tibia and reference lines were placed to bisect the medial border of the distal radial joint surface and parallel to the distal joint surface of the tibia. Scans were acquired using fixed offsets of 9.0mm and 22.0mm proximal from the reference line for the radius and tibia, respectively, as per the standard protocol ^(15)^. One hundred and sixty-eight parallel slices encompassing a 10.197 mm region were acquired at each scan site at the radius and tibia. Scans had a voxel size of 60.7μm. HR-pQCT scans were analyzed using the manufacturer’s standard protocol by one trained individual, which involves slice-matching scans based on a 2D cross-sectional area (Image Processing Language, v5.42, Scanco Medical AG, Wangen-Brüttisellen, Switzerland). Bone variables extracted from standard HR-pQCT analyses included: total, trabecular and cortical vBMD (mg/cm^3^), cortical porosity (%, the segmented pore volume), cortical thickness (mm), trabecular thickness (mm), trabecular separation (mm, the mean space between trabeculae) and trabecular number (1/mm). We also performed finite-element analysis (FEA) at both skeletal sites to estimate bone stiffness (kN/mm) and failure load (kN). We used a 5-point grading scale ^(16)^ to categorise motion artefacts in scans. Scans graded five were excluded from all analyses, scans graded four were excluded from bone microarchitecture and strength analyses, and those graded 3 or lower were included in all analyses. Seven radial scans were given a grade of 4. CVs for all outcomes have been published elsewhere ^(14)^.

Participants attended a Melbourne Pathology clinic and had a 20mL fasting blood sample collected by a qualified technician. Serum samples were assayed for 25(OH)D (Reference #: 310600; DiaSorin Inc., USA), parathyroid hormone (PTH; Reference #: 07251068190; Roche Diagnostics, Australia), type 1 C-telopeptide (CTX; Reference #: 11972308122; Roche Diagnostics, Australia), and type 1 procollagen N-terminal peptide (P1NP; Reference #: 03141071190; Roche Diagnostics, Australia) levels. The CV for 25(OH)D was 9% and ranged between 3.3-5.0% for PTH, 5.6-7.3% for CTX, and 5.7-8.4% for P1NP. Estimated glomerular filtration rate (eGFR) was calculated using serum creatinine (CV: 1.76-2.54%; Reference #: 05168589 190; Roche Diagnostics, Australia), age and sex ^(17)^.

Hand grip strength was measured using a Jamar Plus Digital hydraulic hand grip dynamometer (Patterson Medical, Bolingbrook, IL, USA). In a seated position, participants were instructed to hold the instrument and flex their shoulder until their arm was parallel to the ground at shoulder height, grip and squeeze with the maximum force for 3-5 seconds. Participants performed the test three times with each hand, alternating hands between tests, and were given 60 seconds of rest between each attempt. The maximum value from the six tests was used to calculate hand grip strength (kg) ^(18)^.

Participants were instructed to climb a single flight of 10 steps as quickly as possible, using the handrail for support if preferred ^(19)^. After walking back to the bottom of the staircase, participants were given a 60-second break. The test was repeated twice and the average time taken to climb the flight of steps across both trials was used.

Participants completed the timed up-and-go (TUG) test from a seated position with their backs against the backrest. They were instructed to stand up from a chair, walk to the end of a marked 3-meter course at their usual walking speed, turn around, return to the chair, and sit down ^(20)^. The timer was started as a participant removed their back from the backrest and stopped once the participant had returned to the chair and placed their back against the backrest. The time taken to complete the trial was recorded.

Participants completed the Short Physical Performance Battery (SPPB), a validated tool for measuring physical performance and disability in older adults ^(21)^. It consisted of three tasks; a chair stand test (also known as a 5-time sit-to-stand test), standing balance tests, and a gait speed test. A score between 0-12 was calculated based on performance in these tests and higher scores indicate better physical performance. Participants started the chair stand test from a standing position and were instructed to cross their arms touching opposite shoulders and attempt to sit and stand as quickly as possible five times without stopping, finishing in a seated position. Study staff started the clock as soon as they told participants to begin the test; they counted repetitions with participants and stopped the clock once the fifth repetition was completed (placed their back against the backrest). Tests were terminated if participants were unable to complete all five repetitions, used their arms for assistance, or the timer passed one minute. Scores between 0-4 were noted based on the time taken to complete five stands. Standing balance tests included three variations with different difficulty; semi-tandem, full-tandem and side-by-side standing. When performing semi-tandem stands, participants were instructed to place the heel of one foot alongside the big toe of the other foot and time in this position for 10 seconds. If completed successfully, participants were instructed to place their preferred foot directly in front of the other, touching heel-to-toes to complete a full-tandem stand and they were instructed to hold the position for ten seconds. If participants could not hold the semi-tandem stand for ten seconds, they were instructed to stand with their feet touching side-by-side and hold this position for ten seconds. Following completion of the balance tests, a score of 0–4 was assigned based on performance in these tests. Gait speed was measured using a 2.44 m course and a score of 0–4 was assigned based on the time it took to walk this distance at a normal walking speed.

Lower limb muscle power was estimated from the maximal vertical jump test on a force platform (Leonardo Mechanograph; Novotec Medical GmbH). Participants performed three maximal vertical jump trials without arm swing, with 60-second rest intervals between trials. Vertical ground reaction forces were calculated from the point of stationary standing to the point of landing to determine maximal vertical jump power (W), which was normalised to participant body weight (kg), using the manufacturer’s standard software (Version 4.2). The maximum value out of all three attempts was used in analyses.

We defined adherence as the percentage of attended sessions out of a total of 34 weekly sessions (the 8-month intervention period included approximately 34-35 sessions per participant); attendance data was recorded by OsteoStrong^®^. An adverse event (AE) was defined as any untoward medical occurrence in a participant during the study period, irrespective of whether it was related to study activities. Furthermore, a serious AE (SAE) was defined as any untoward medical occurrence that resulted in death, life-threatening event, disability or incapacitation, or required hospitalisation (minimum 1 overnight stay) at a hospital or emergency ward for observation and/or treatment that would not have otherwise been appropriate in a physician’s office or outpatient treatment. Participants were provided with contact details of investigators and instructed to report any AE and/or SAE that occurred during the intervention period. AEs or SAEs that occurred during or shortly after an OsteoStrong^®^ session were evaluated by the research team to be likely or possibly due to the intervention.

### 2.13 Statistical Analysis

Although the primary outcomes of interest were related to feasibility and safety, the sample size calculation was based on an estimate of a 4% improvement in total hip aBMD following 8 months of OsteoStrong^®^. This anticipated improvement was a conservative estimate, lower than what has been reported elsewhere ^(3)^, in response to the OsteoStrong^®^ intervention. A sample size of 44 (accounting for a 20% loss to follow-up) was deemed sufficient to detect a 4% mean difference in total hip aBMD from baseline to 8 months, with a type-II error rate of 5% and >80% power.

Normality assessments were performed for continuous data using boxplots and Shapiro– Wilk tests. Descriptive statistics were calculated as means ± standard deviation (SD), frequencies (percentages), or as median (interquartile range [IQR]) for non-parametric data. Our main analyses followed intention-to-treat principles. Baseline and follow-up values are presented as estimated marginal means with 95% confidence intervals (95%CI). Eight-month changes in all outcome measures are presented as mean change (95%CI). Mean changes were analysed with linear mixed models using an unstructured covariance matrix and all models included the respective outcome measure as a dependent variable and time (2 levels). We performed per-protocol analyses in participants with at least 6 months (≥26 weeks) of adherence. We also performed sensitivity analyses in participants who achieved OsteoStrong^®^’s specified osteogenic thresholds for upper, lower, core and exercises in ≥26 sessions. In order to reach an osteogenic threshold, participants needed to produce force equivalent to 2.5 multiples of bodyweight (MOB) for the upper growth trigger, 4.2 MOB for the lower growth trigger, 1.5 MOB for the core growth trigger and 2.5 MOB for the postural growth trigger. MOB was estimated via the Spectrum^®^ system. Individual and mean percentage change in DXA- and HR-pQCT-determined densitometric estimates were plotted with least significant change (LSC; i.e. the smallest difference between two sequential measurements that can confidently be considered true biological change rather than a product of measurement error or variability) values calculated based on precision data published elsewhere ^(14)^. For two participants, baseline CTX values were below the assay’s lower limit of quantification (<70 ng/L). These values were conservatively imputed as 70 ng/L for the primary analysis. Sensitivity analyses excluding these datapoints were conducted to assess the robustness of the findings. For all analyses, a P-value of <0.05 or 95% confidence interval not including the null point was considered statistically significant. All data were analysed using SPSS Statistics Version 24 (IBM, USA).

## 3.0 Results

A total of 769 participants were screened for this study to achieve the target sample size of n=44 (conversion rate 5.7%), with common exclusion reasons including regularly engaging in habitual physical activity likely to improve bone health (e.g., resistance training), having normal aBMD, and the use of anti-osteoporosis medication (Figure 1). The cohort of 44 postmenopausal women had a mean age of 61.2 ± 5 years and 81% reported having completed university or higher education (Table 1). Based on baseline DXA measurements, most women had osteopenia (73%), the remainder (27%) had osteoporosis, and 70% self-reported a previous (lifetime) fracture. Mean protein intake was >1.3g/kg/day and three women consumed less than 0.75g/kg/day. Five women had 25OHD levels <50nmol/L (3/5 had levels <30nmol/L).

**Figure 1.**
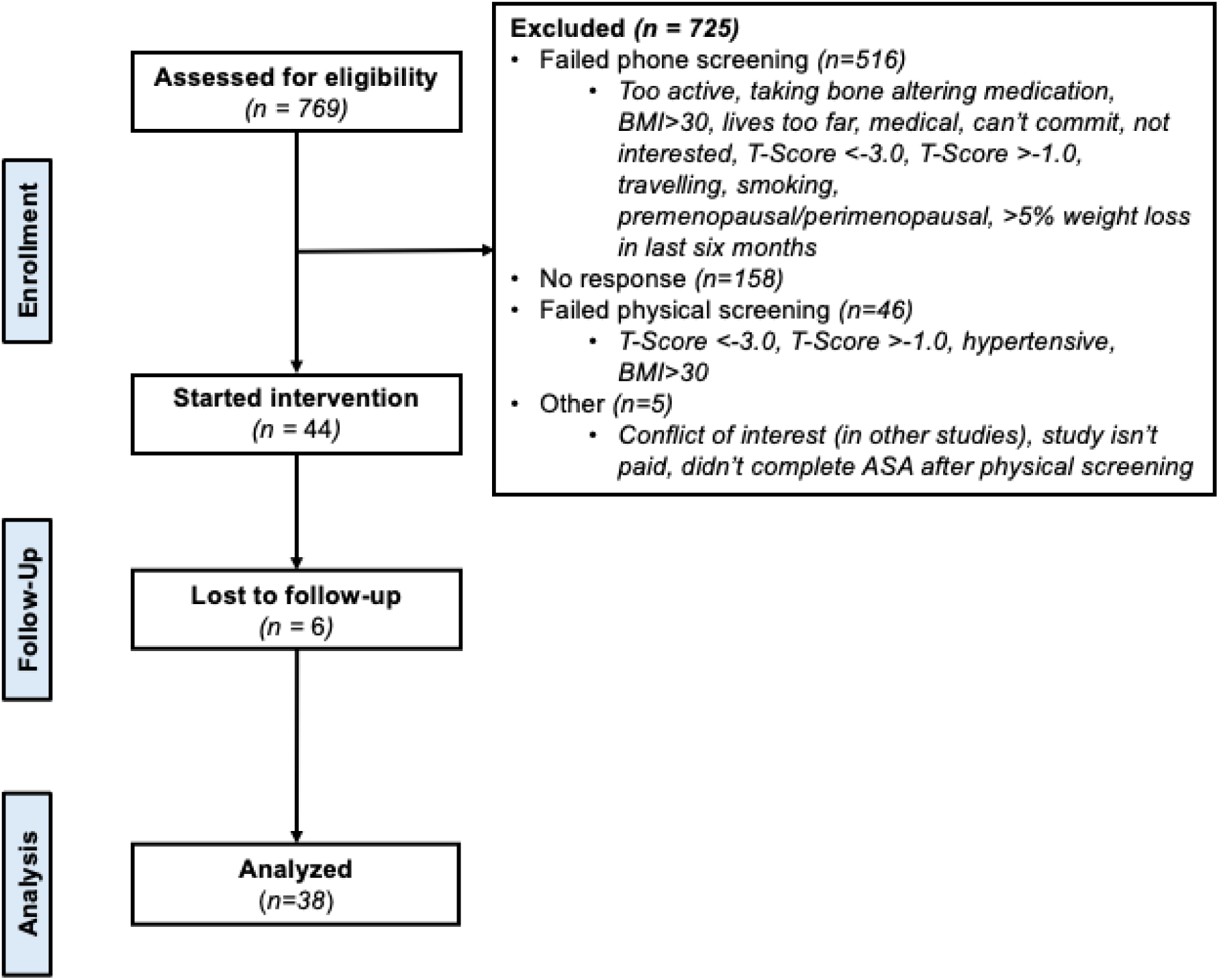
Flow of participants

**Table 1.**
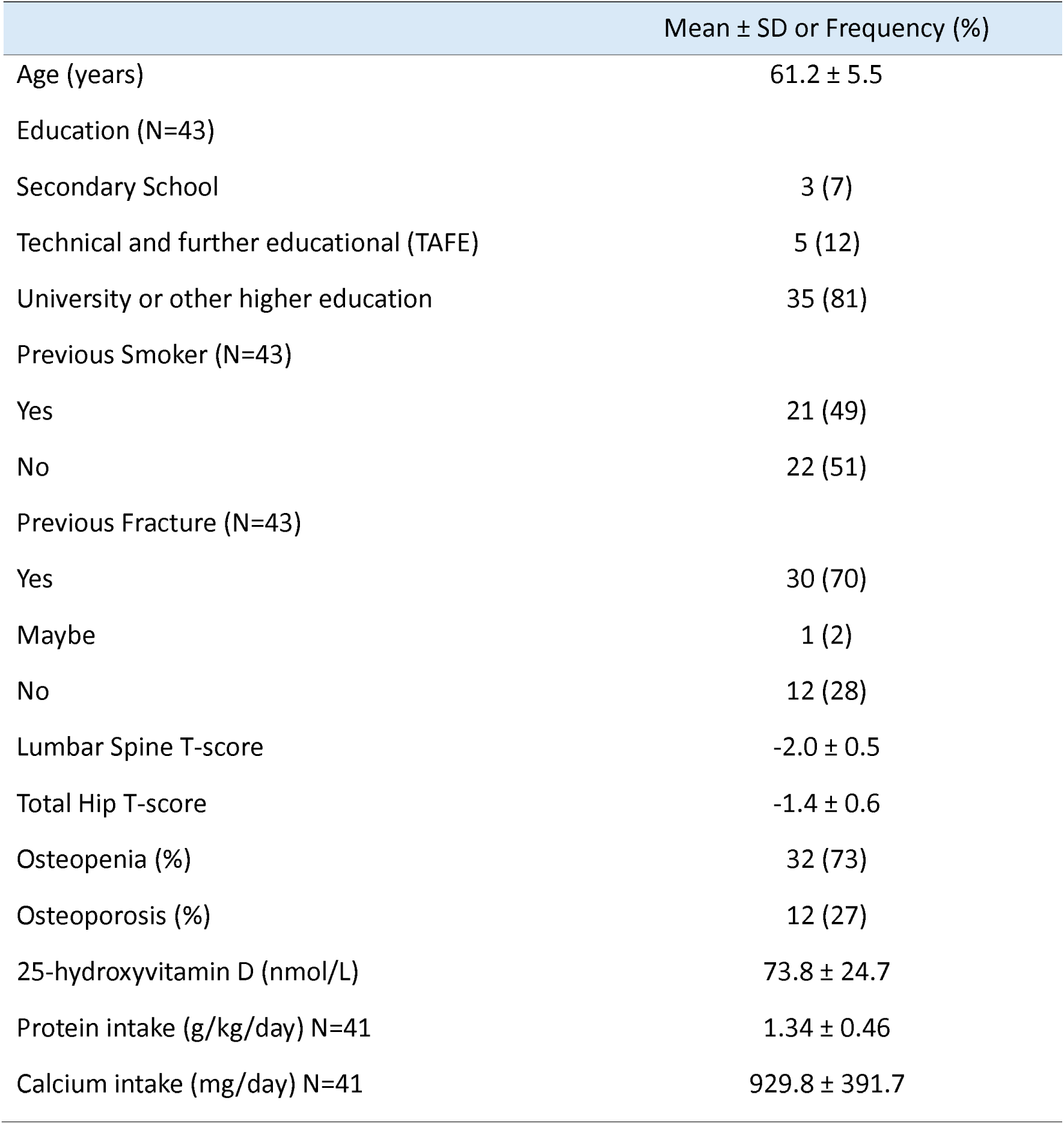
Descriptive Characteristics.

Six participants withdrew or were lost to follow-up during the 8-month intervention. Reasons for withdrawal or loss to follow-up included moving interstate (n=4) and personal reasons (n = 2), which resulted in an attrition rate of 14%. Mean adherence to the intervention was 83 ± 28% in the entire cohort and 93 ± 9% in the 38 participants who completed the intervention. A total of four adverse events were recorded (Supplemental Table S1). One participant experienced a flare-up of a pre-existing medical condition after an OsteoStrong^®^ session (likely intervention-related). Another reported Achilles tendonitis, which developed soon after baseline testing (not intervention-related). A third participant fractured her forearm after falling off a bicycle (not intervention-related), and the fourth suffered a sacroiliac strain during the intervention (possibly intervention-related). Three of the four participants who reported an adverse event continued exercising throughout the study (adherence: >97%); the individual who sustained a forearm fracture stopped attending OsteoStrong^®^ sessions after the injury at week 21 (adherence: 65%) but completed follow-up testing.

Tables 2-4 present changes in bone variables estimated by DXA, HR-pQCT and pQCT after 8 months. TBS decreased (-1.8%) but there were no changes in total hip, femoral neck, or lumbar spine aBMD. At the distal radius, we observed a decrease in total (-1.8%), trabecular (-1.7%) and cortical (-0.8%) vBMD and cortical thickness (-0.8%). At the distal tibia, cortical vBMD decreased (-0.7%) and trabecular separation increased (0.8%). All other estimates of bone density, microarchitecture and strength did not change by 8 months. Individual changes in DXA- and HR-pQCT-determined densitometric estimates plotted against LSCs at various skeletal sites are presented in Supplemental Figure S1. Most individual changes were below the LSC at all skeletal sites.

**Table 2.** Changes in DXA-estimated bone variables following 8 months of OsteoStrong^®^.

|  | Baseline | Follow-Up | Mean Change | P-Value |
| --- | --- | --- | --- | --- |
| Femoral Neck aBMD (g/cm<sup>2</sup>) | 0.642 (0.623, 0.662) | 0.643 (0.622, 0.663) | 0.001 (-0.007, 0.008) | 0.972 |
| Total Hip aBMD (g/cm<sup>2</sup>) | 0.774 (0.751, 0.797) | 0.771 (0.747, 0.795) | -0.003 (-0.007, 0.002) | 0.262 |
| Lumbar Spine aBMD (g/cm<sup>2</sup>) | 0.823 (0.805, 0.841) | 0.825 (0.805, 0.845) | 0.002 (-0.005, 0.009) | 0.575 |
| TBS (L1-L4) | 1.302 (1.284, 1.319) | 1.278 (1.257, 1.298) | -0.024 (-0.039, -0.009) | 0.003 |

Data are mean (95% confidence intervals); aBMD – areal bone mineral density; TBS – trabecular bone score

**Table 3.** Changes in HRpQCT-estimated bone variables at the radius following 8 months of OsteoStrong^®^.

|  | Baseline | Follow-Up | Mean Change | P-Value |
| --- | --- | --- | --- | --- |
| Total vBMD (mg HA/cm<sup>3</sup>) | 256.5 (241.0, 271.9) | 251.8 (236.4, 267.2) | -4.6 (-6.2, -3.1) | <0.001 |
| Trabecular vBMD (mg HA/cm<sup>3</sup>) | 117.8 (107.0, 128.6) | 115.8 (105.3, 126.3) | -2.0 (-2.8, -1.1) | <0.001 |
| Cortical vBMD (mg HA/cm<sup>3</sup>) | 828.6 (812.2, 844.9) | 821.9 (803.7, 840.0) | -6.7 (-11.5, -1.9) | 0.007 |
| Trabecular number (1/mm) | 1.201 (1.131, 1.272) | 1.193 (1.122, 1.265) | -0.008 (-0.018, 0.002) | 0.118 |
| Trabecular thickness (mm) | 0.220 (0.217, 0.223) | 0.220 (0.216, 0.223) | 0.000 (-0.001, 0.001) | 0.922 |
| Trabecular separation (mm) | 0.838 (0.779, 0.898) | 0.849 (0.784, 0.914) | 0.011 (-0.001, 0.021) | 0.061 |
| Cortical Porosity (%) | 0.007 (0.006, 0.009) | 0.007 (0.006, 0.009) | 0.000 (-0.001, 0.001) | 0.979 |
| Cortical Thickness (mm) | 0.864 (0.819, 0.909) | 0.857 (0.811, 0.903) | -0.007 (-0.012, -0.002) | 0.008 |
| Stiffness (kN/mm) | 46094 (43734, 48454) | 45788 (43226, 48350) | -306 (-1258, 646) | 0.516 |
| Failure Load (kN) | -2488 (-2617, -2359) | -2471 (-2612, -2330) | 17 (-39, 73) | 0.537 |

Data are mean (95% confidence intervals); vBMD – volumetric bone mineral density

**Table 4.** Changes in HRpQCT-estimated bone variables at the tibia following 8 months of OsteoStrong^®^.

|  | Baseline | Follow-Up | Mean Change | P-Value |
| --- | --- | --- | --- | --- |
| Total vBMD (mg HA/cm<sup>3</sup>) | 240.5 (226.6, 254.4) | 239.5 (225.7, 253.3) | -1.0 (-2.1, 0.03) | 0.056 |
| Trabecular vBMD (mg HA/cm<sup>3</sup>) | 138.3 (127.1, 149.5) | 137.8 (126.7, 149.0) | -0.5 (-1.1, 0.1) | 0.098 |
| Cortical vBMD (mg HA/cm<sup>3</sup>) | 794.4 (775.8, 813.1) | 788.5 (769.4, 807.5) | -6.0 (-8.0, -3.9) | <0.001 |
| Trabecular number (1/mm) | 1.183 (1.11, 1.255) | 1.177 (1.103, 1.25) | -0.006 (-0.018, 0.007) | 0.376 |
| Trabecular thickness (mm) | 0.247 (0.242, 0.252) | 0.247 (0.243, 0.252) | 0.000 (-0.001, 0.001) | 0.609 |
| Trabecular separation (mm) | 0.893 (0.758, 1.028) | 0.900 (0.763, 1.036) | 0.007 (0.001, 0.012) | 0.021 |
| Cortical Porosity (%) | 0.034 (0.03, 0.038) | 0.035 (0.031, 0.039) | 0.001 (-0.001, 0.003) | 0.164 |
| Cortical Thickness (mm) | 1.224 (1.154, 1.294) | 1.229 (1.160, 1.298) | 0.005 (-0.006, 0.016) | 0.366 |
| Stiffness (kN/mm) | 140971 (134366, 147577) | 140721 (133911, 147532) | -250 (-1735, 1235) | 0.740 |
| Failure Load (kN) | -7700 (-8043, -7358) | -7689 (-8041, -7336) | 11 (-61, 84) | 0.744 |
| **Proximal Site 66% (pQCT)** |  |  |  |  |
| Cortical vBMD (mg/cm<sup>3</sup>) | 1087 (1077, 1096) | 1087 (1078, 1096) | 0 (-3, 4) | 0.824 |
| SSI Polar (mm<sup>3</sup>) | 1996 (1905, 2087) | 2011 (1903, 2119) | 15 (-38, 70) | 0.559 |

Data are mean (95% confidence intervals); vBMD – volumetric bone mineral density; SSI -strength-strain index

Changes in physical activity and function after 8 months of OsteoStrong^®^ are presented in Table 5. Stair climb times and chair stand times decreased (-2.6% and -10.4%, respectively) and SPPB scores increased (1.7%); 95% of participants achieved a perfect score in the balance component of the SPPB at baseline, while 100% achieved a perfect score at follow-up. There were no changes in hand grip strength or any other physical function outcomes. Blood biochemistry and body composition changes are reported in Tables 6 and 7. There were no changes in eGFR, PTH, CTX or P1NP at 8 months (Table 6), nor were there any changes in fat mass, ALM, visceral adipose tissue or calf muscle density (Table 7).

**Table 5.**
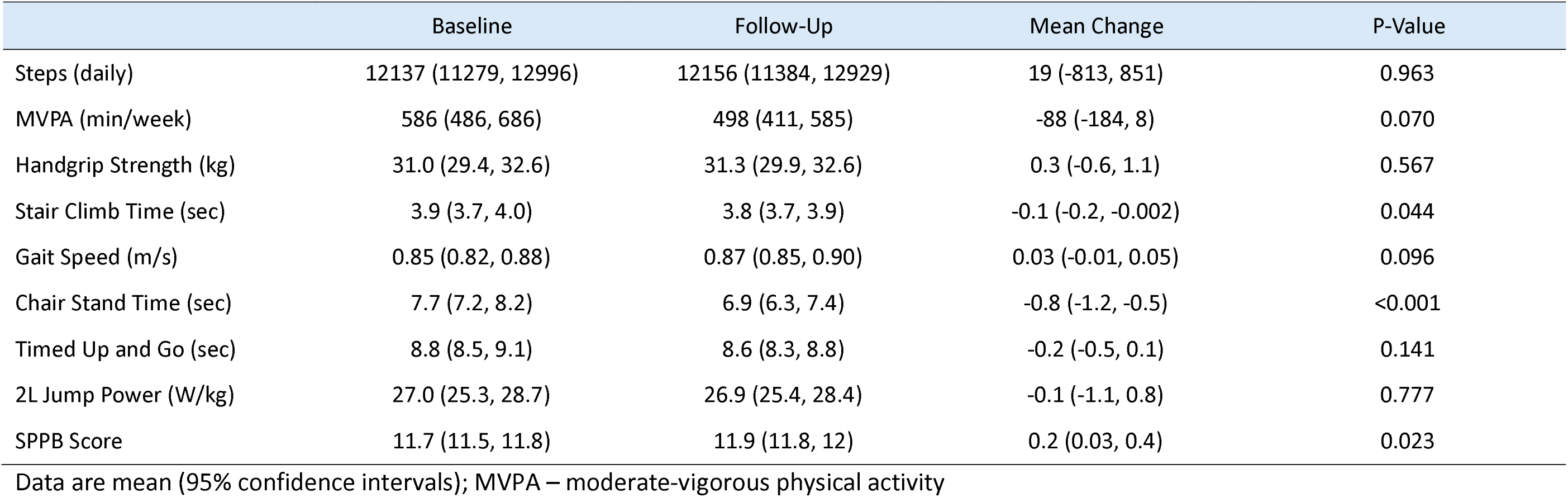
Changes in physical activity and function following 8 months of OsteoStrong^®^.

**Table 6.**
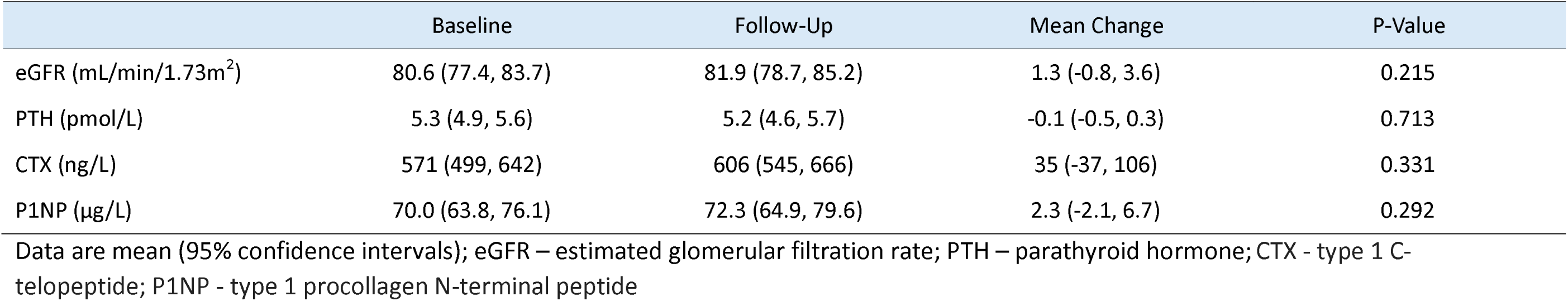
Blood biochemistry changes following 8 months of OsteoStrong^®^.

**Table 7.**
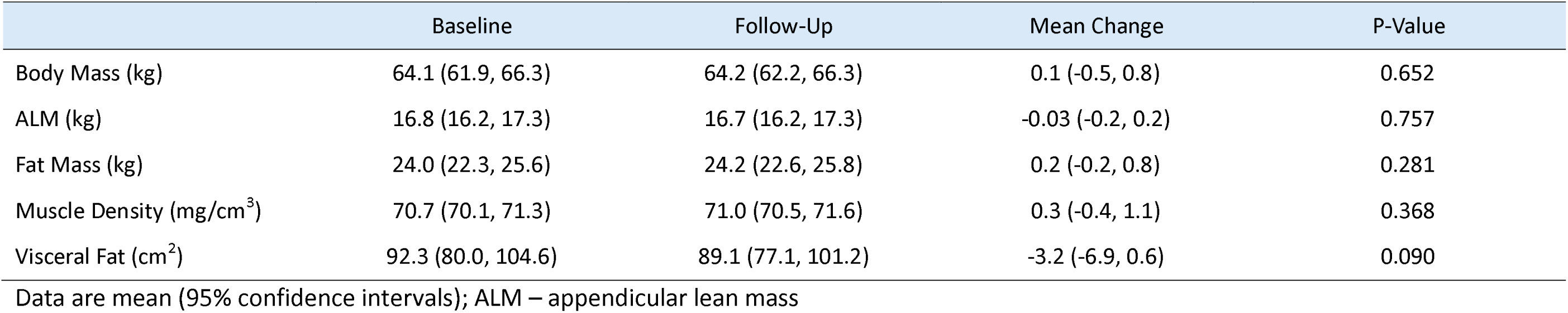
Body Composition changes following 8 months of OsteoStrong^®^.

We performed per-protocol analyses among 36 participants who attended OsteoStrong^®^ for ≥6 months (75% of prescribed sessions) (Supplementary Tables S2-S7). All results remained unchanged except for the improvement in stair climb times and SPPB scores, which became non-significant. We also performed sensitivity analyses in subgroups of participants who reached the OsteoStrong^®^-determined osteogenic threshold in ≥26 sessions (equivalent to a 6-month cumulative exercise dose). In women who achieved their lower growth trigger threshold (n=30), all results remained unchanged except total vBMD (mean change: - 1.137mgHA/cm³ [95% CI: -2.042, -0.231]) and trabecular vBMD (mean change: - 0.657mgHA/cm³ [95% CI: -1.302, -0.011]) at the distal tibia decreased. Similarly, in women who reached their postural growth trigger threshold (n=32), the increase in trabecular separation at the distal tibia also became non-significant (mean change: 0.006mm [95% CI: - 0.0004, 0.012]). No changes were observed in those who achieved their core growth trigger threshold (n=31), and an insufficient number of participants achieved their upper growth trigger threshold (n=9) to perform meaningful analyses. Sensitivity analyses omitting two baseline CTX values <70 ng/L did not affect the main or per-protocol analyses.

## 4.0 Discussion

This 8-month single-arm pilot study demonstrated that OsteoStrong^®^ was feasible based on ≥80% attendance rates and few adverse events, and also improved some measures of physical function, in postmenopausal women with low BMD . However, Osteostrong^®^ did not improve bone density, microarchitecture or strength, nor did it influence biochemical markers of bone health, or body composition. While the lack of a no-treatment control group in our study precludes conclusions on whether this intervention can preserve bone health over time, these data do not indicate any improvement in bone density or strength in this population participating in OsteoStrong^®^ for 8 months.

Adherence to Osteostrong^®^ was high (83 ± 28% overall; 93 ± 9% in those who completed the study). While there is no consensus on acceptable exercise adherence, rates ≥80% are usually considered satisfactory ^(22)^. Similar results were reported in LIFTMOR-M, which utilised bioDensity™ machines (78.5 ± 14.8%) ^(4)^. The only study that has evaluated the Spectrum^®^ system did not report exercise session adherence ^(5)^. Attrition rates of 14% were similar to what would be expected in exercise studies ^(23)^. High adherence and low attrition may be attributed to the fact that the program was supervised, as well as the minimal weekly time commitment required of participants (a single 10-15-minute session per week) ^(24–26)^. Participants were also able to engage with the intervention at no financial cost, which may have influenced long-term adherence. There were no AEs deemed intervention-related and two minor AEs that were possibly intervention-related. However, it should be noted that participants were expected to self-report AEs, which may have led to underreporting, and we did not screen for asymptomatic AEs, such as osteoporotic vertebral fractures. Future studies should monitor for these events while also considering feasibility of sustained engagement for participants following the paid membership model.

For all estimates of bone health, we observed either no significant changes or significant declines, following 8 months of OsteoStrong^®^. No changes were observed in DXA-determined total hip, femoral neck or lumbar spine aBMD, while TBS decreased. Age-related aBMD losses in postmenopausal women are approximately 0.5-2.5% annually, with the greatest declines observed in the early postmenopausal years ^(27,28)^. Thus, the maintenance of aBMD observed in our cohort may suggest a protective effect, but this is speculative given the lack of a control group and that the trial was only 8 months in duration.

As previously noted, the 4% improvement in total hip aBMD our study was powered to detect was conservative relative to 12-month treatment effects claimed by OsteoStrong^®^ (14%) ^(1)^.

For context, RCTs of bisphosphonates typically report aBMD increases of approximately 2- 5% (2-3% total hip; up to 5% lumbar spine) ^(29–31)^, while bone anabolic agents can lead to gains of 4-13% (4-7% total hip; up to 13% lumbar spine) over 12 months ^(29,32)^. Most exercise interventions involving resistance and/or impact training report more modest gains, generally in the range of 1–3% per year at the total hip and <5% at the lumbar spine ^(4,33–35)^. In the LIFTMOR-M trial, 8-month changes in total hip, femoral neck, and lumbar spine aBMD were <2% in the bioDensity™ group, although greater gains at the lumbar spine were observed in response to high-intensity resistance and impact training (4.1%) ^(4)^. In the present study, 8 months of OsteoStrong^®^ resulted in total hip, femoral neck and lumbar spine aBMD changes of <0.01%.

This is the first study of OsteoStrong^®^ to evaluate changes in bone microarchitecture and strength. At the distal radius, total, trabecular and cortical vBMD and cortical thickness decreased. At the distal tibia, cortical vBMD decreased and trabecular separation increased. The deterioration we observed in the abovementioned densitometric and microarchitecture estimates was similar to annual mean changes reported elsewhere in postmenopausal women ^(36)^. Furthermore, decreases in total vBMD at the radius exceeded the LSC ^(14)^, suggesting this was a meaningful change. The lack of improvement and deterioration in bone health in our study may be partially attributed to the low training volume and frequency (15 minutes, once weekly) in OsteoStrong^®^. A secondary analysis of an 18-month RCT in 172 middle-aged and older men demonstrated that both training frequency and volume were significant predictors of aBMD change at the femoral neck and lumbar spine, with those who performed ≥2 sessions per week experiencing the greatest improvements ^(37)^. Systematic reviews and meta-analyses of controlled trials generally support these findings, with higher frequency, volume and intensity leading to the greatest improvements in bone health ^(38–40)^. Notably, many effective bone-targeted exercise interventions also include a high impact component such as jumping, hopping, or drop landings ^(33,41)^, which is absent in OsteoStrong^®^.

OsteoStrong^®^ improved stair climb and chair stand time and SPPB scores. Chair stand time improved by approximately 10% (mean change: -0.8 seconds), a result consistent with findings from the LIFTMOR-M trial, which reported a 4.5 ± 1.7% improvement ^(4)^. Improvements in chair stand time likely contributed to the increase in SPPB scores, given that no changes in gait speed were observed and most women (≥95%) had the highest possible scores in the balance component at both time points. Similarly, stair climb time improved by 2% (mean change: -0.1 seconds), which is considered an indirect measure of lower-limb muscle power, however, this was not paralleled by changes in two-leg jump power assessed using jumping mechanography. We also observed no changes in hand grip strength during this intervention. Collectively, these findings suggest OsteoStrong^®^ might enhance lower-limb muscle function, which could translate into reduced fall risk in older adults ^(42,43)^.

OsteoStrong^®^ had no effect on biochemical or body composition outcomes. These findings are consistent with a recent OsteoStrong^®^ trial that similarly reported no changes in CTX or P1NP after 12 months ^(5)^. While visceral fat decreased in our study, this did not achieve statistical significance and the lack of effect on ALM, total body or fat mass, and muscle density is unsurprising given the low training frequency and volume of OsteoStrong^®^, which is not targeted at improving body composition per se.

This study had several strengths. This was the first study to (1) assess the feasibility of the OsteoStrong^®^ in postmenopausal women with low bone density, (2) explore how it influences bone microarchitecture and strength (determined by HR-pQCT), body composition (determined by DXA and pQCT), physical function and biochemical markers of kidney and parathyroid function, and (3) take into account OsteoStrong^®^-generated force data (measured via the Spectrum^®^ system) to explore whether participants who reached their specified ‘osteogenic threshold’ during sessions had more pronounced changes in bone outcomes. Other strengths of this study include the reporting of precision data for all bone variables and the 8-month duration of the study, which provided sufficient time to observe exercise-related bone adaptations and allowed for comparability with other bone-targeted interventions ^(4,35,44)^.

Our study also had limitations. The absence of a control group limited our ability to account for potential confounders such as time (i.e., ageing) and repeated exposure to testing (i.e., learning effects). Additionally, except for total hip aBMD, changes in BMD and bone microarchitecture observed in this study should be considered exploratory given our limited statistical power. No adjustments were made for multiple comparisons, and our target population consisted exclusively of postmenopausal women with low BMD who were rigorously screened to exclude those engaging in activities or presenting with conditions known to affect bone and mineral metabolism. While this enhanced internal validity by reducing confounding, it may limit the applicability of our results to more heterogeneous populations (such as those with normal BMD or comorbidities) commonly seen in real-world settings. Furthermore, the provision of free memberships to OsteoStrong^®^ for all participants may have positively influenced adherence and so pragmatic trials may be required to evaluate the feasibility and cost-effectiveness of this intervention.

In conclusion, 8 months of OsteoStrong^®^ was a feasible and well-tolerated intervention for postmenopausal women with low BMD, demonstrated by good adherence, few adverse events, and low attrition. Although some improvements in physical function were observed, bone outcomes, including total hip aBMD, either remained stable or declined over the intervention period. No changes were detected in biochemical markers of bone metabolism or body composition. These findings suggest that while OsteoStrong^®^ may enhance aspects of physical function, it is unlikely to be effective as a standalone intervention for improving bone health in this population.

## Data Availability

Most data in this manuscript collected by Monash University, as well as Spectrum data provided by OsteoStrong, are available in online supplementary material. Any additional data presented are available upon reasonable request to the authors.

## Acknowledgements

We would like to thank all the participants involved in the study, and the contributions of OsteoStrong^®^ staff and Mx Cat Shore-Lorenti.

## Australian New Zealand Clinical Trials Registration

ACTRN12620000224921p

## Data availability

Data in this manuscript collected by Monash University, as well as Spectrum^®^ data provided by OsteoStrong^®^, are available in online supplementary material.

All authors have read and approved the manuscript.

## SUPPLEMENTARY DATA

**Supplemental Figure S1.**
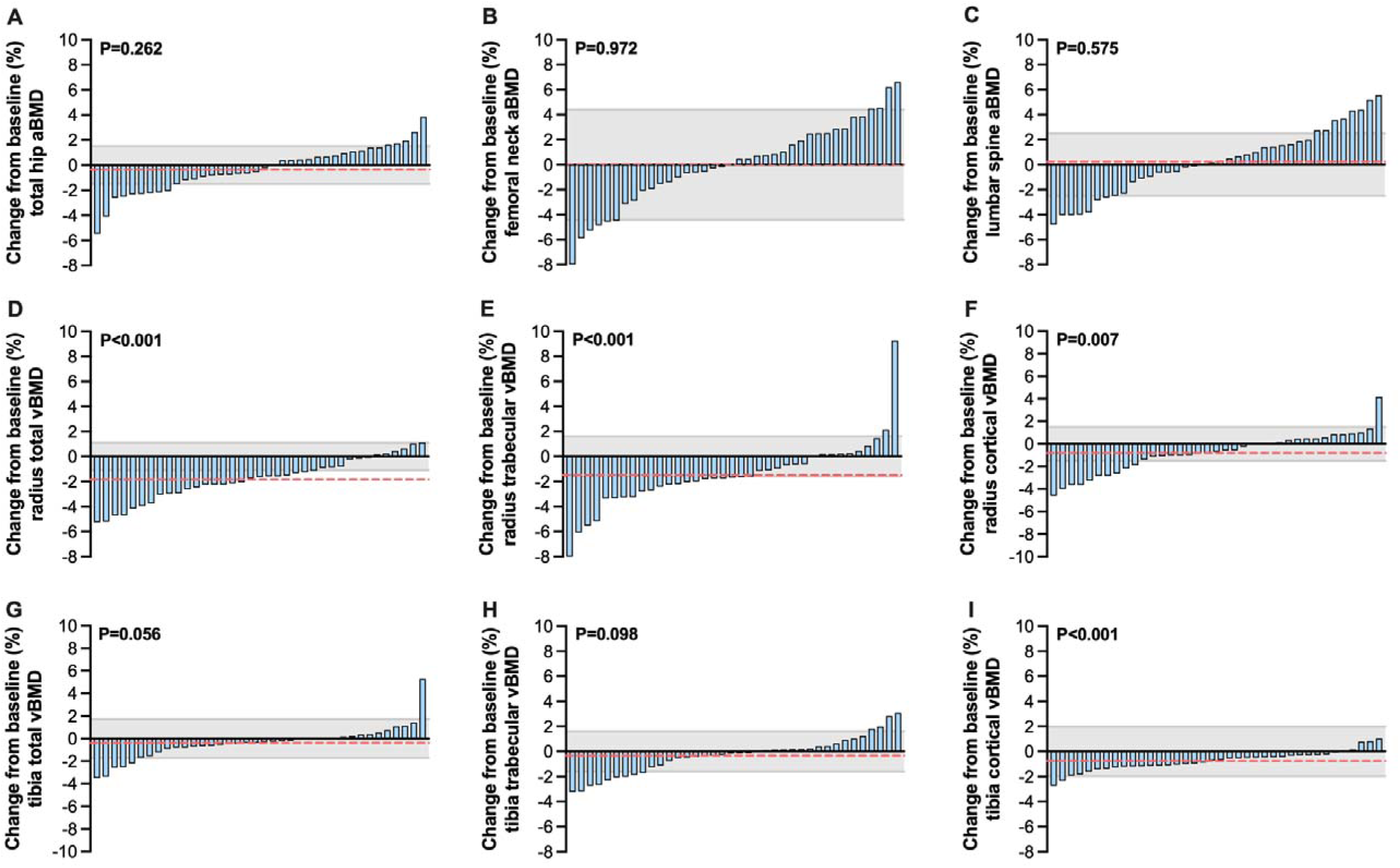
Individual (blue bars), mean (dotted orange line) and least significant (shaded grey) changes in DXA- and HR-pQCT-determined densitometric estimates at various skeletal sites.

**Table S1.**
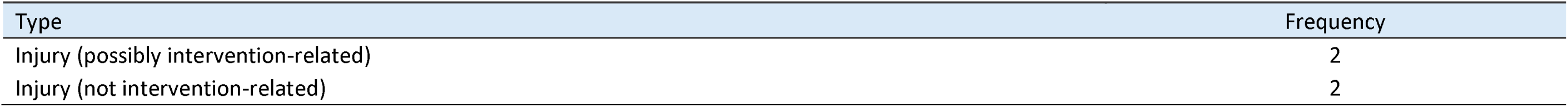
List of related, potentially related and unrelated adverse events reported by study participants.

| Type | Frequency |
| --- | --- |
| Injury (possibly intervention-related) | 2 |
| Injury (not intervention-related) | 2 |

**Table S2.** Per protocol analyses of changes in DXA-estimated bone variables following 8 months of OsteoStrong^®^.

|  | Baseline | Follow-Up | Mean Change | P-Value |
| --- | --- | --- | --- | --- |
| Femoral Neck aBMD (g/cm<sup>2</sup>) | 0.635 (0.614, 0.657) | 0.634 (0.612, 0.656) | -0.001 (-0.009, 0.005) | 0.585 |
| Total Hip aBMD (g/cm<sup>2</sup>) | 0.769 (0.742, 0.795) | 0.766 (0.738, 0.793) | -0.003 (-0.008, 0.002) | 0.192 |
| Lumbar Spine aBMD (g/cm<sup>2</sup>) | 0.818 (0.798, 0.838) | 0.819 (0.797, 0.84) | 0.001 (-0.007, 0.009) | 0.774 |
| TBS (L1-L4) | 1.295 (1.274, 1.315) | 1.269 (1.249, 1.290) | -0.025 (-0.041, -0.010) | 0.002 |

Data are mean (95% confidence intervals); N=38; aBMD – areal bone mineral density; TBS – trabecular bone score

**Table S3.** Per protocol analyses of changes in HR-pQCT-estimated bone variables at the radius following 8 months of OsteoStrong^®^.

|  | Baseline | Follow-Up | Mean Change | P-Value |
| --- | --- | --- | --- | --- |
| Total vBMD (mg HA/cm<sup>3</sup>) | 254.5 (236.5, 272.4) | 250.2 (232.3, 268.2) | -4.2 (-5.8, -2.7) | <0.001 |
| Trabecular vBMD (mg HA/cm<sup>3</sup>) | 114.3 (102.3, 126.3) | 112.6 (100.8, 124.3) | -1.8 (-2.6, -0.9) | <0.001 |
| Cortical vBMD (mg HA/cm<sup>3</sup>) | 829.7 (811.0, 848.3) | 823.0 (802.5, 843.6) | -6.6 (-11.7, -1.6) | 0.012 |
| Trabecular number (1/mm) | 1.175 (1.097, 1.254) | 1.168 (1.088, 1.249) | -0.007 (-0.017, 0.004) | 0.201 |
| Trabecular thickness (mm) | 0.220 (0.216, 0.224) | 0.220 (0.216, 0.224) | 0.000 (-0.001, 0.001) | 0.877 |
| Trabecular separation (mm) | 0.861 (0.793, 0.928) | 0.872 (0.797, 0.946) | 0.011 (-0.001, 0.022) | 0.069 |
| Cortical Porosity (%) | 0.007 (0.006, 0.009) | 0.007 (0.006, 0.009) | 0.000 (-0.001, 0.001) | 0.945 |
| Cortical Thickness (mm) | 0.867 (0.814, 0.921) | 0.862 (0.808, 0.917) | -0.005 (-0.010, 0.0004) | 0.033 |
| Stiffness (kN/mm) | 45882 (43173, 48592) | 45688 (42737, 48639) | -194 (-1205, 817) | 0.697 |
| Failure Load (kN) | -2470 (-2618, -2322) | -2460 (-2623, -2298) | 10 (-50, 69) | 0.749 |

Data are mean (95% confidence intervals); N=38; vBMD – volumetric bone mineral density

**Table S4.** Per protocol analyses of changes in HR-pQCT-estimated bone variables at the tibia following 8 months of OsteoStrong^®^.

|  | Baseline | Follow-Up | Mean Change | P-Value |
| --- | --- | --- | --- | --- |
| Total vBMD (mg HA/cm<sup>3</sup>) | 237.9 (222.1, 253.6) | 237.0 (221.3, 252.7) | -0.9 (-1.9, 0.2) | 0.098 |
| Trabecular vBMD (mg HA/cm<sup>3</sup>) | 135.7 (123.4, 147.9) | 135.1 (122.9, 147.4) | -0.5 (-1.2, 0.7) | 0.079 |
| Cortical vBMD (mg HA/cm<sup>3</sup>) | 793.2 (772.4, 813.9) | 787.4 (766.2, 808.6) | -5.8 (-7.9, -3.7) | <0.001 |
| Trabecular number (1/mm) | 1.149 (1.073, 1.225) | 1.143 (1.066, 1.221) | -0.006 (-0.019, 0.008) | 0.387 |
| Trabecular thickness (mm) | 0.247 (0.242, 0.253) | 0.248 (0.243, 0.253) | 0.001 (-0.001, 0.003) | 0.795 |
| Trabecular separation (mm) | 0.925 (0.762, 1.087) | 0.932 (0.768, 1.096) | 0.007 (0.001, 0.013) | 0.020 |
| Cortical Porosity (%) | 0.035 (0.030, 0.039) | 0.036 (0.031, 0.04) | 0.001 (-0.001, 0.003) | 0.239 |
| Cortical Thickness (mm) | 1.221 (1.143, 1.300) | 1.228 (1.150, 1.306) | 0.006 (-0.005, 0.018) | 0.282 |
| Stiffness (kN/mm) | 139975 (132454, 147496) | 139782 (132029, 147535) | -193 (-1756, 1370) | 0.804 |
| Failure Load (kN) | -7639 (-8030, -7249) | -7630 (-8032, -7228) | 9 (-67, 86) | 0.804 |
| **Proximal Site 66% (pQCT)** |  |  |  |  |
| Cortical vBMD (mg/cm<sup>3</sup>) | 1084 (1073, 1094) | 1085 (1075, 1096) | 2 (-1, 4) | 0.174 |
| SSI Polar (mm<sup>3</sup>) | 1997 (1899, 2094) | 2013 (1900, 2127) | 17 (-40, 74) | 0.553 |

Data are mean (95% confidence intervals); N=38; vBMD – volumetric bone mineral density; SSI -strength-strain index

**Table S5.** Per protocol analyses of changes in physical activity and function following 8 months of OsteoStrong^®^.

|  | Baseline | Follow-Up | Mean Change | P-Value |
| --- | --- | --- | --- | --- |
| Steps (daily) | 12010 (11112, 12907) | 12002 (11209, 12795) | -8 (-882, 867) | 0.986 |
| MVPA (min/week) | 555 (452, 659) | 483 (390, 577) | -72 (-173, 29) | 0.157 |
| Handgrip Strength (kg) | 31.2 (29.4, 33.0) | 31.5 (30.0, 33.0) | 0.3 (-0.5, 1.2) | 0.475 |
| Stair Climb Time (sec) | 3.8 (3.7, 4.0) | 3.8 (3.6, 3.9) | -0.1 (-0.1, 0.01) | 0.127 |
| Gait Speed (m/s) | 0.85 (0.82, 0.88) | 0.87 (0.84, 0.90) | 0.03 (-0.01, 0.06) | 0.119 |
| Chair Stand Time (sec) | 7.7 (7.2, 8.2) | 6.8 (6.3, 7.4) | -1.0 (-1.2, -0.4) | <0.001 |
| Timed Up and Go (sec) | 8.7 (8.4, 9.1) | 8.6 (8.3, 8.8) | -0.2 (-0.5, 0.1) | 0.256 |
| 2L Jump Power (W/kg) | 27.0 (25.1, 28.9) | 26.9 (25.3, 28.5) | -0.1 (-1.0, 0.8) | 0.832 |
| SPPB Score | 11.7 (11.5, 11.8) | 11.9 (11.7, 12.0) | 0.2 (-0.001, 0.4) | 0.051 |

Data are mean (95% confidence intervals); N=38; MVPA – moderate-vigorous physical activity

**Table S6.** Per protocol analyses of blood biochemistry changes following 8 months of OsteoStrong^®^.

|  | Baseline | Follow-Up | Mean Change | P-Value |
| --- | --- | --- | --- | --- |
| eGFR (mL/min/1.73m<sup>2</sup>) | 81.3 (77.9, 84.6) | 82.0 (78.6, 85.4) | 0.7 (-1.3, 2.8) | 0.474 |
| PTH (pmol/L) | 5.3 (4.9, 5.8) | 5.2 (4.7, 5.8) | -0.1 (-0.5, 0.3) | 0.660 |
| CTX (ng/L) | 549 (469, 629) | 599 (534, 665) | 51 (-26, 128) | 0.189 |
| P1NP (µg/L) | 68.3 (61.8, 74.8) | 70.7 (62.7, 78.8) | 2.5 (-2.1, 7.0) | 0.278 |

Data are mean (95% confidence intervals); N=38; eGFR – estimated glomerular filtration rate; PTH – parathyroid hormone; CTX - type 1 C-telopeptide; P1NP - type 1 procollagen N-terminal peptide

**Table S7.** Per protocol analyses of body composition changes following 8 months of OsteoStrong^®^.

|  | Baseline | Follow-Up | Mean Change | P-Value |
| --- | --- | --- | --- | --- |
| Body Mass (kg) | 63.9 (61.5, 66.3) | 64.1 (61.9, 66.4) | 0.2 (-0.5, 0.9) | 0.508 |
| ALM (kg) | 16.8 (16.1, 17.4) | 16.8 (16.1, 17.4) | 0.0 (-0.2, 0.2) | 0.864 |
| Fat Mass (kg) | 23.8 (22.0, 25.5) | 24.1 (22.4, 25.8) | 0.3 (-0.2, 0.9) | 0.206 |
| Muscle Density (mg/cm<sup>3</sup>) | 70.5 (69.8, 71.2) | 71.0 (70.4, 71.6) | 0.5 (-0.3, 1.3) | 0.203 |
| Visceral Fat (cm<sup>2</sup>) | 91.2 (77.6, 104.9) | 88.1 (74.9, 101.4) | -3.1 (-7.0, 0.8) | 0.114 |

Data are mean (95% confidence intervals); N=38; ALM – appendicular lean mass

